# Nowcasting and Forecasting the Spread of COVID-19 in Iran

**DOI:** 10.1101/2020.04.22.20076281

**Authors:** Hamidreza Masjedi, Jomar F. Rabajante, Fatemeh Bahranizadd, Mohammad Hosein Zare

**Affiliations:** Department of Medical Physics, School of Medicine, Shahid Sadoughi University of Medical Sciences and Health Services, Yazd, Iran; Institute of Mathematical Sciences and Physics, University of the Philippines Los Baños, Laguna 4031 Philippines; Faculty of Education, University of the Philippines Open University, Laguna 4031 Philippines

## Abstract

**Introduction:** As of early December 2019, COVID-19, a disease induced by SARS-COV-2, has started spreading, originated in Wuhan, China, and now on, have infected more than 2 million individuals throughout the world.

**Purpose:** This study aimed to nowcast the COVID-19 outbreak throughout Iran and to forecast the trends of the disease spreading in the upcoming month.

**Methods:** The cumulative incidence and fatality data were extracted from official reports of the National Ministry of Health and Medical Educations of Iran. To formulate the outbreak dynamics, six phenomenological models, as well as a modified mechanistic Susciptible-Exposed-Infectious-Recovered (SEIR) model, were implemented. The models were calibrated with the integrated data, and trends of the epidemic in Iran was then forecasted for the next month.

**Results:** The final outbreak size calculated by the best fitted phenomenological models was estimated to be in the range of 68,486 to 118,923 cases; however, the calibrated SEIR model estimated that the outbreak would rage again, starting from April 26. Moreover, projected by the mechanistic model, approximately half of the infections have undergone undetected.

**Conclusion:** Although the advanced phenomenological models perfectly fitted the data, they are incapable of applying behavioral aspects of the outbreak and hence, are not reliable enough for authorities’ decision adoptions. In contrast, the mechanistic SEIR model alarms that the COVID-19 outbreak in Iran may peak for the second time, consequent to lifting the control measures. This implies that the government may implement a more granular decision making to control the outbreak.

## 1 Introduction

COVID-19 is a coronavirus disease caused by the virus SARS-CoV-2. The outbreak of this disease was first observed in Wuhan, China, around December 2019 (1). As of 16 April 2020, more than 2 million confirmed cases were reported worldwide, with more than 130 thousand deaths. In Iran, more than 76 thousand infected individuals were reported in which 6.25% of these were expired cases (2).

One of the basic parameters in studying the epidemics of COVID-19 is knowing the basic reproductive number *R*_0_ (3). This number provides a measure of how contagious the disease is. *R*_0_ is the average number of individuals in a fully susceptible population that a primary case can directly infect during the whole infectiousness period. There are many studies estimating the *R*_0_ for Iran. Chen et al. (2020) estimated that the *R*_0_ as of 31 March 2020 is 4.51 (4). Fadaei and Rahmani (2020) estimated that *R*_0_ is about 4.7, and as of 3 April 2020, the effective *R* had been reduced to below 2 (5).

There are reports that several neighboring countries had imported COVID-19 cases from Iran. It was estimated that the ascertainment rate in Iran is around 0.6% as of 25 February 2020 (6). Shortage in and delay in the provision of medical, pharmaceutical, and laboratory equipment contribute to the adverse impact of the pandemic on the communities in Iran (7). A modeling paper had suggested that it is essential to forecast the timing of the epidemic peak, and to estimate the healthcare, government and public readiness for “flattening the curve”, such as by promoting social distancing (8). It was also suggested that measures should still be in place to hinder a second wave of the outbreak (9, 10).

In this paper, the dynamics of the outbreak in Iran was modeled using phenomenological (data-fitting) and mechanistic (SEIR) approaches. SEIR, which stands for Susceptible-Exposed-Infectious-Recovered, has been modified to include possible asymptomatics and the isolation of infected individuals (11, 12).

## 2 Methods

### 2.1 Data

The daily updates of officially reported COVID-19 cases were obtained from the website of the National Ministry of Health and Medical Educations of Iran (13). The data includes the number of laboratory-confirmed case incidences as well as fatalities from 19 February 2020 to 16 April 2020. In order to avoid the irregularities and reporting lags affecting short time-series (14, 15), all the models were fashioned for the cumulative data. Figure 1 illustrates the cumulative number of cases and deaths as a function of time.

**Figure 1.**
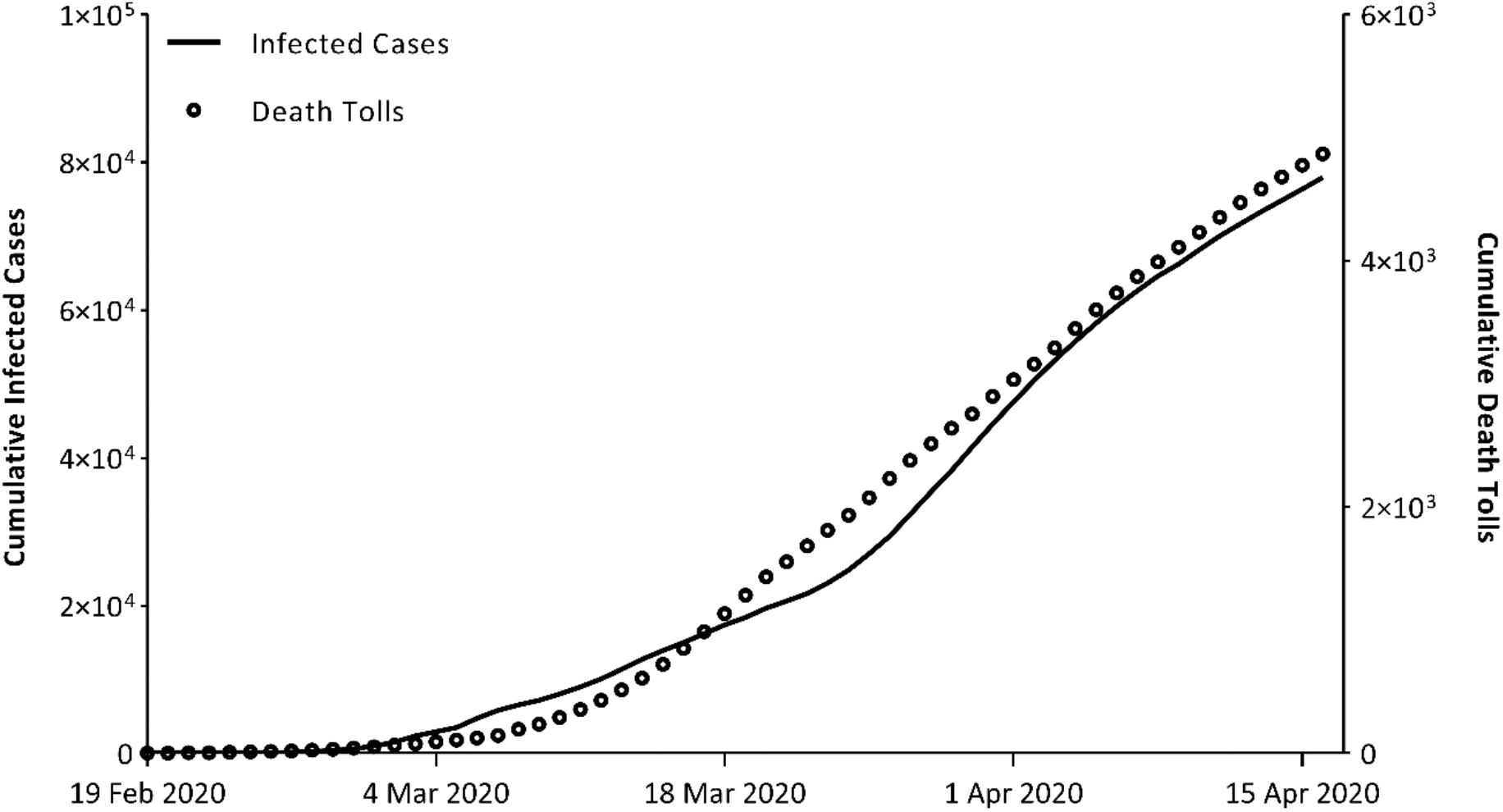
Cumulative number of COVID-19 cases and death tolls in Iran from 19 February to 15 April 2020

### 2.2 Phenomenological modeling

#### 2.2.1 Models

The generalized-Richards model (GRM) (16) was implemented to characterize the disease transmissibility and generate forecasts on the epidemic in Iran. This model extends the simple Richards model (17) by introducing *p*, a parameter that decelerates the growth speed.

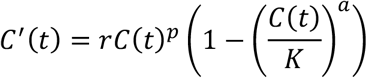

Here, *C(t)* is the cumulative number of cases at time *t, r* is the growth rate, *K* represents the final size of the epidemic, and *a* is a measure of deviation from the S-shaped logistic curve.

For comparison, the five well-known phenomenological models, including Richards model (RM), generalized-logistic growth model (GLGM), logistic growth model (LGM), generalized-growth model (GGM), and exponential growth model (EGM), were also calibrated to the data. A concise description of each model and the comparisons are provided in the supplementary material.

#### 2.2.2 Model fitting

The models were calibrated based on the daily number of cumulative cases using MATLAB (The Mathworks, Inc.). The MATLAB built-in least-squares fitting function (*lsqcurvefit)* was used by calling the *trust-region reflective* algorithm to implement the fitting process.

#### 2.2.3 Uncertainty of estimation

In order to derive the uncertainty of parameter estimations, a parametrized bootstrap approach was followed (18). In this context, by assuming a Poisson distribution for the error structure around the best-fit solution, M bootstrap datasets were generated, and then, by refitting the model to each, the parameters were derived. Following the simulations, the 95% confidence interval of parameter estimations was considered as the 2.5th to 97.5th percentile. A comprehensive description of the approach is given by Chowell (19).

### 2.3 Mechanistic modeling

#### 2.3.1 Model structure

Figure 2 portrayed a schematic diagram of the mechanistic model incorporated in this study for estimating the current situation of the disease as well as forecasting the foreseeable future of the COVID-19 outbreak in Iran. Standing on the traditional SEIR model (20), our model considers four main stocks representing population classes: susceptible (S), exposed (E), infectious (I), and recovered (R). Despite the SEIR model supposition of the perfect mixing of the whole population, this assumption is violated in the real world. Hence, to account for this discrepancy, the initial number of susceptible population (*S*_*0*_) was calculated by the following equation, proposed by Hartfield and Alizon (21) for the final size of an epidemic:

**Figure 2.**
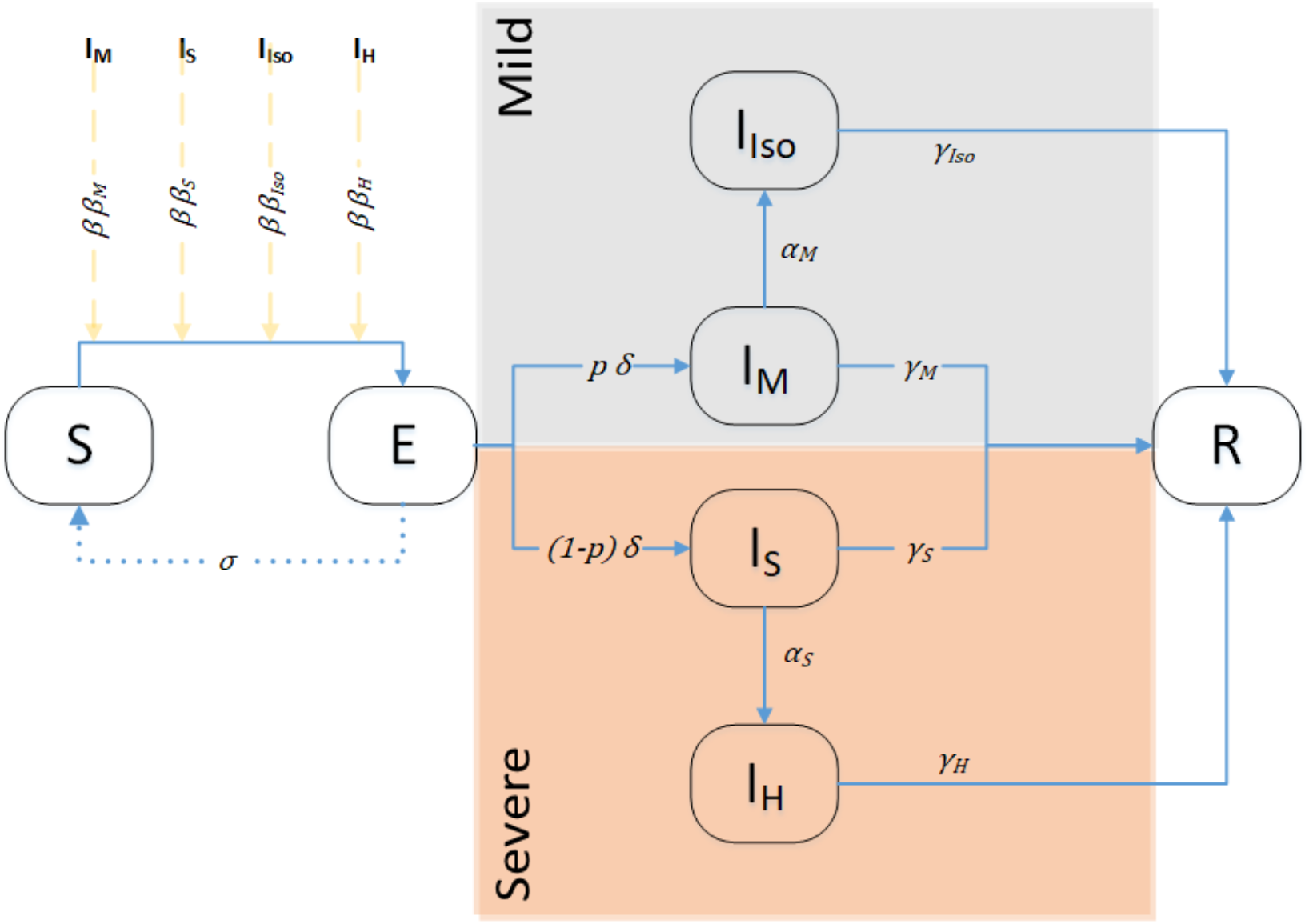
A schematic diagram of the compartmental model adopted to simulate the spread of the COVID-19 in Iran; S: susceptible, E: exposed, mildly infected (I_M_), isolated mildly infected (I_Iso_), severely infected (I_S_), and hospitalized (I_H_), R: recovered

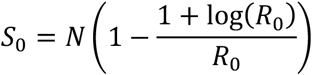

where *N* represents the population size, and *R*_*0*_ accounts for the basic reproductive number.

The connections across the compartments were also depicted in Figure 2. As of this writing, since the outbreak is still in progress, to estimate the number of beds required and other crucial statistics, the infectious class was categorized into four subgroups, including mildly infected (I_M_), isolated mildly infected (I_Iso_), severely infected (I_S_), and hospitalized (I_H_). A reinforcing loop was then embedded in the model that regulates the spread of COVID-19 from infectious to susceptible populations. For the purpose of taking the infectiousness of each subgroup into account, the transmissibility of each infectious subgroup was weighted by a factor denoted by *β*_*i*_, where the subscription i refers to each infectious subgroup. The *β*_*S*_ related to the severely infected was considered as unity, and the infectiousness of the other infectious subgroups was scaled relative to the severely infected compartment.

#### 2.3.2 Influence of control measures

To reflect the influence of control measures (e.g., school and university closures, and travel restrictions) imposed by the government, it was assumed that the transmission rate *R*(*t*) decays exponentially as a function of time (22) with a decay constant 4, after a threshold time, *T*_*0*_. In view of the fact that all the control measures were not simultaneously applied, *T*_*0*_ was determined through scenario simulation. Hence, the *R*(*t*) was defined by the following equation:

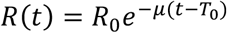

Moreover, to address the effect of protection actions such as washing hands, a new parameter, σ was defined that represents the transition rate from the exposed compartment back to the susceptible class. This parameter was estimated through the calibration of the model, and multiple scenarios were then generated by considering different values of σ (subsection 2.3.5).

#### 2.3.3 Model parameters

A brief definition of the parameters used to regulate disease transmission and individual transitions across the compartments are given in Table 1. Note that all the parameters were derived through the model calibration with real cumulative incidence data. For the calculation of the transmission rate (*β*), the basic reproductive number was extracted by forming the next-generation matrix (23), and then, the expression was solved for *β*. The instruction is given in the supplementary material.

**Table 1.** A concise definition of the parameters and their values.

| Parameter | Definition |
| --- | --- |
| $R_0$ | Basic reproductive number |
| $\beta$ | Transmission rate |
| $\beta_S$ | Infectiousness of severely infected (considered as unity to scale the level of infectiveness of other infectious subgroups) |
| $\beta_M$ | Relative infectiousness of mildly infected to severely infected compartment |
| $\beta_{Iso}$ | Relative infectiousness of isolated mildly infected to severely infected compartment |
| $\beta_H$ | Relative infectiousness of hospitalized to severely infected compartment |
| $\delta_M$ | Transition rate from exposed class to mildly infected subgroup |
| $\delta_S$ | Transition rate from exposed class to severely infected subgroup |
| $p$ | Proportion of mildly infected population to the total infected cases |
| $\alpha_M$ | Isolation rate of mildly infected cases |
| $\alpha_S$ | Hospitalization rate of severely infected cases |
| $\gamma_M$ | Recovery rate of mildly infected cases |
| $\gamma_S$ | Recovery rate of severely infected cases |
| $\gamma_{Iso}$ | Recovery rate of isolated mildly infected cases |
| $\gamma_H$ | Recovery rate of hospitalized |
| $T_0$ | Threshold time at which control measures were applied |
| $\mu$ | Decay constant |
| $\sigma$ | Protection rate of exposed people from being infectious |

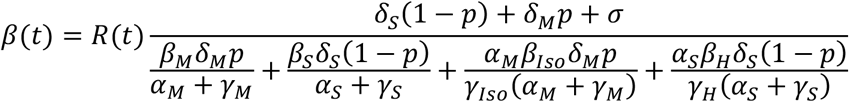

#### 2.3.4 Simulation

The simulations were performed by a house-made code using MATLAB. The code was designed to numerically solve the system of ordinary differential equations (odes) and extract the model uncertainty. The parametrized bootstrap approach, explained in subsection 2.2.3, was followed by MATLAB.

#### 2.3.5 Scenarios

The National authorities had started lifting the control measures as of April 11, 2020 (24). In order to analyze the consequences of this, considering that the reproductive number may increase exponentially through time since April 11 with the same constant, 4, multiple scenarios were assessed by adjusting various levels for protection rate, σ. The σ was set to the fitted value and ±20%, ±50%, ±100%, and +200% tolerances were assumed. Based on each scenario, a forecast with a 95% prediction interval was constructed for the next month of the COVID-19 outbreak in Iran.

### 2.4 Death tolls

The National Ministry of Health and Medical Educations publishes publically-accessible brief reports on the situation of COVID-19 in Iran and the world on a daily basis, including the current status of the COVID-19, patients epidemiological characteristics, results of simulations, etc. (25). By extracting the disease-induced fatality rate from the reports, categorized by sex and age groups, and applying them to the Iran age and sex structure (26), the age-and sex-adjusted fatality rate was calculated for Iran as well as for each province separately. The death tolls were then estimated through multiplying the estimated total number of infections from the mechanistic model by the fatality rate.

## 3 Results

### 3.1 Data-fitting

In total, six phenomenological models involving generalized-Richards model, Richards model, generalized-logistic growth model, logistic growth model, generalized-growth model, and exponential growth model were fitted to the cumulative number of cases. Figure 3 illustrates the best-fit curves to the data given the six models. The estimated parameters, as well as the parameters describing fitting accuracy for each model, are given in the supplementary material (Tables S.1, S.2).

**Figure 3.**
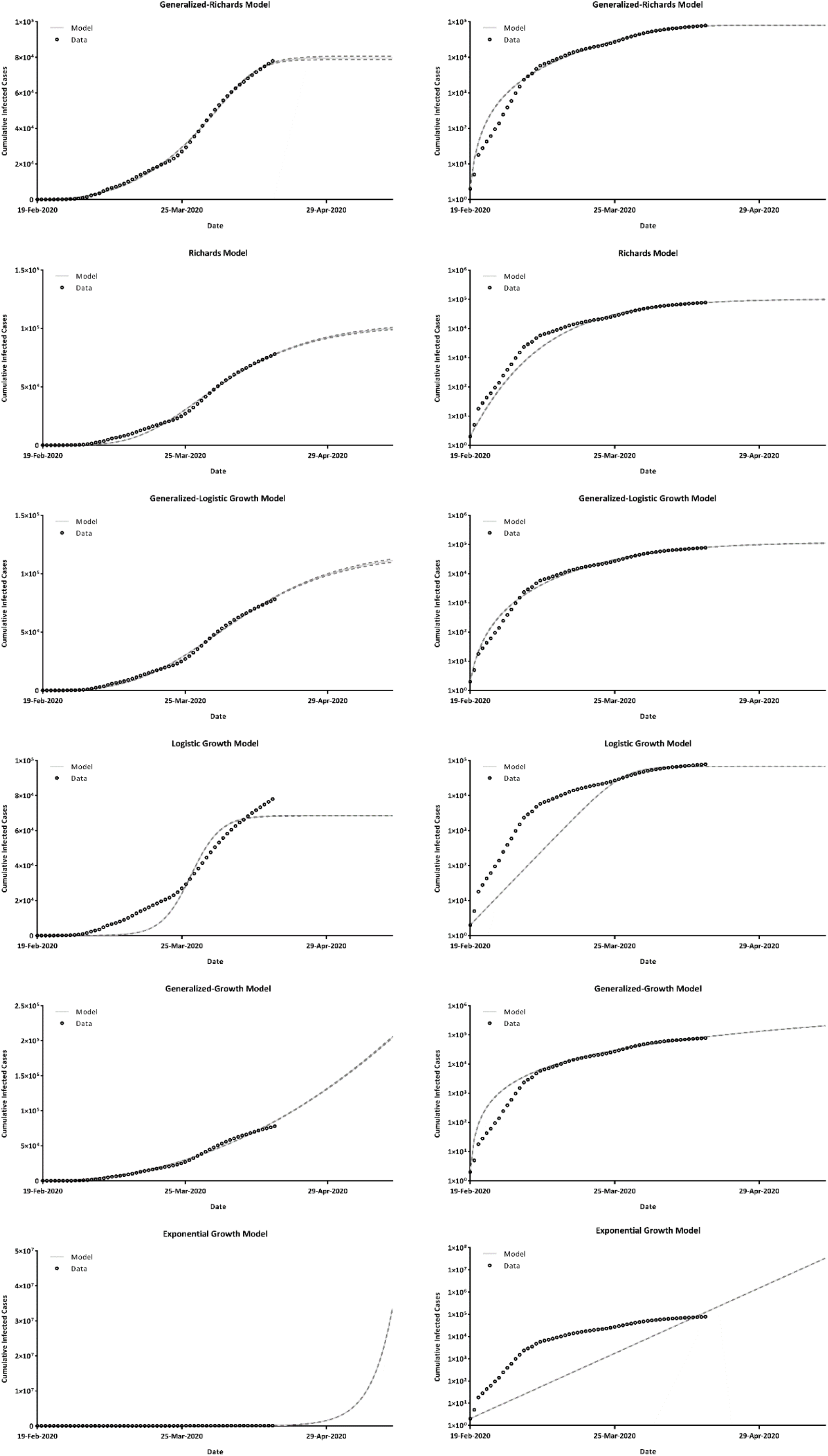
The best-fit curves to the cumulative infected cases against time for six models, including generalized-Richards model, Richards model, generalized-logistic growth model, logistic growth model, generalized-growth model, and exponential growth model. The dashed lines represent the 95% prediction interval. The graphs on the left are on a linear scale, and the right-side graphs are on the semi-log scale.

In light of the results, the GRM estimated the epidemic size to be 79,681. In parallel, Richards model, generalized-logistic growth model, and logistic growth model projected 10,4482, 118,923, and 68,486 infected cases at the final state of the outbreak in Iran. Regarding the goodness of fittings, the generalized-logistic growth model exhibited the lowest mean absolute percentage error (MAPE=22%), whereas the generalized-Richards model showed the lowest root mean squared error (RMSE=1010) and mean absolute error (MAE=810).

### 3.2 Mechanistic modeling

#### 3.2.1 Baseline SEIR model

Figure 4 represents the results of the simulation for the baseline SEIR model. Demonstrated by Figure 4, the findings suggesting that another outbreak will start around April 26, consequent to lifting the quarantine measures. Moreover, as shown in Figure 4.E, the model predicts that the total number of infected cases with COVID-19 is roughly 50% higher comparing to reported numbers, implying that a bunch of cases is not detected. The estimated parameters, as well as the goodness of fit, are reported in the supplementary material (Tables S.3, S.4).

**Figure 4.**
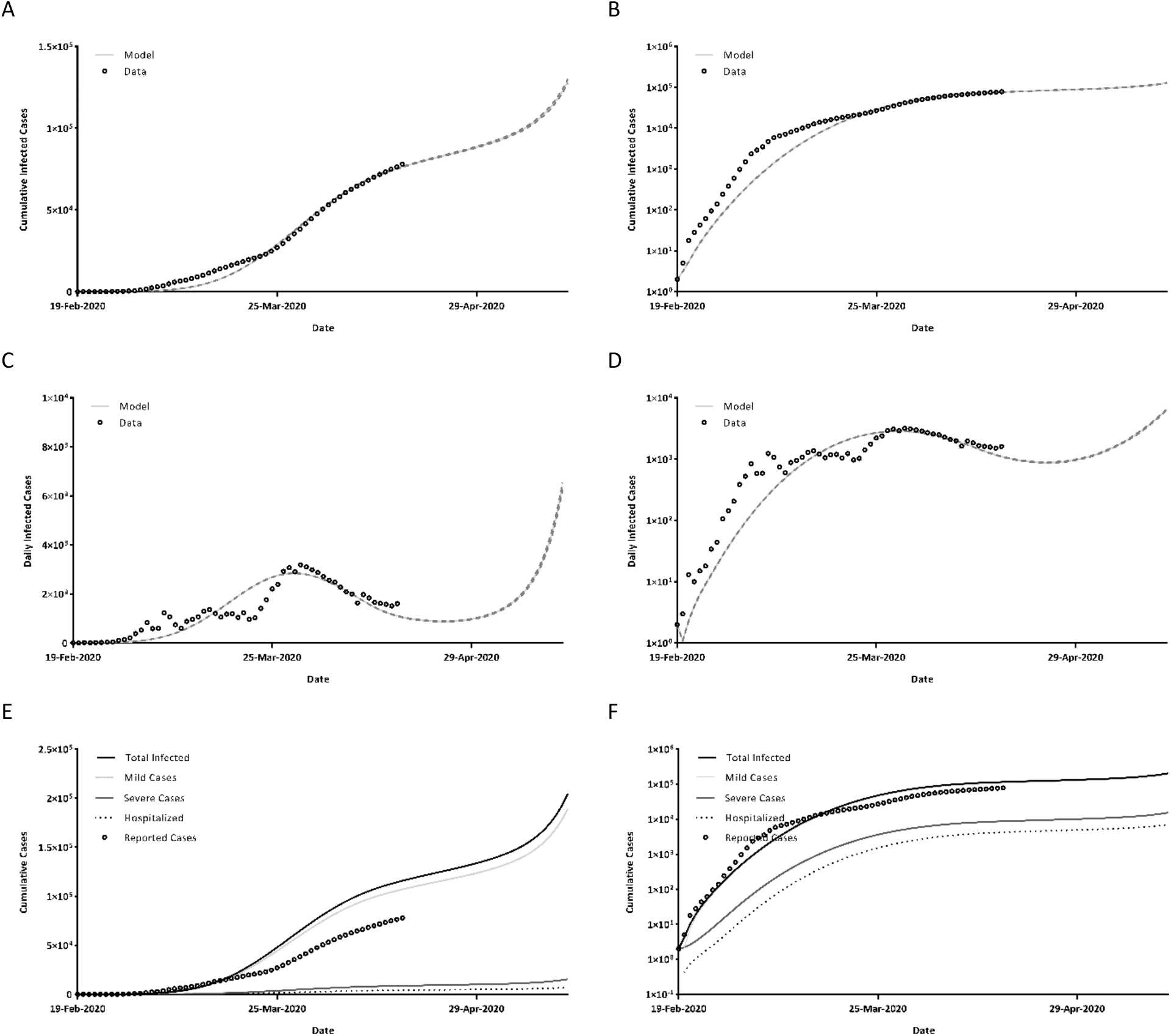
Simulation-based estimation of a, b) cumulative reported cases, c, d) daily reported cases, and e, f) cumulative number of total infected cases, severe cases, mild cases as well as hospitalized cases, based on the ground SEIR model. The graphs on the left are on a linear scale, and the right-side graphs are on the semi-log scale.

Under the baseline scenario, assuming that the epidemic has not been controlled, it is estimated that as of mid-May, around 220,000 cases would be infected, 7.5% of whom get severely infectious, and nearly 7,500 of them need hospitalization. Note that the number of infected cases tends to increase as long as no effective actions will be put into practice, and hence, there will be no termination of the epidemic.

Figure 5 shows the time-evolution of the basic reproductive number. As evidenced, the epidemic raged with an *R*_*0*_ of 3.66 for a week, then, contracted below unity after March 27 and started raising from April 11, as the quarantine measures started to lift, and passed unity after April 23.

**Figure 5.**
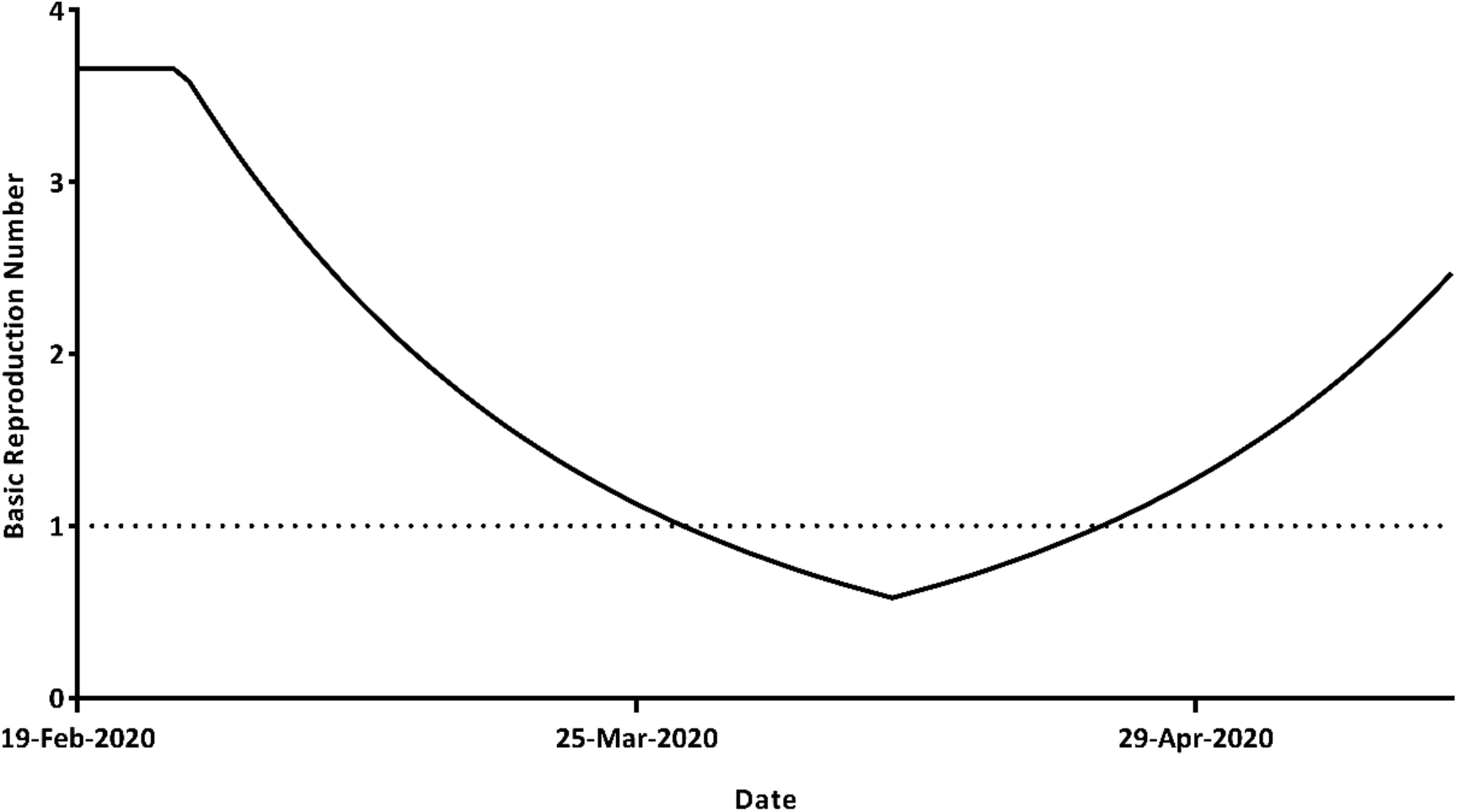
Variation of basic reproductive number as a function of time; The dashed line highlights the R_0_=1

#### 3.2.2 Scenarios

Described in subsection 2.3.5, by tilting the value of protection rate (σ), multiple scenarios were generated. Figure 6 illustrates the total number of infected cases, severe cases, mild cases, and hospitalized cases for each scenario. Expectedly, by increasing the σ, the number of infected cases would decrease and vice versa.

**Figure 6.**
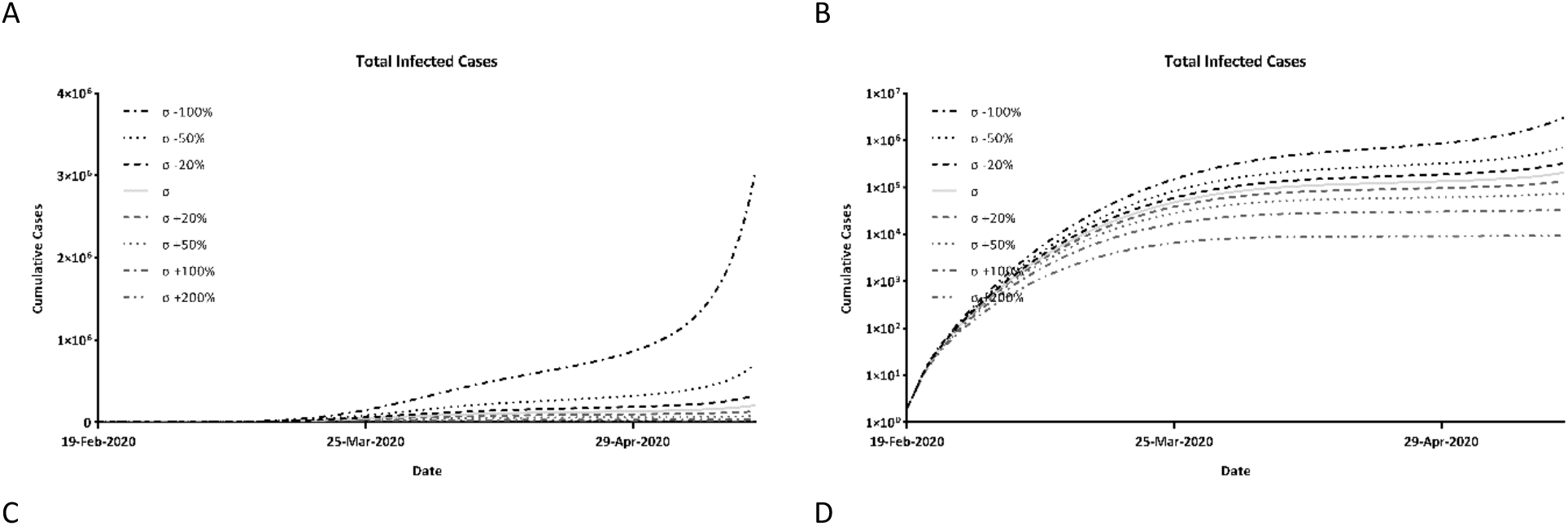
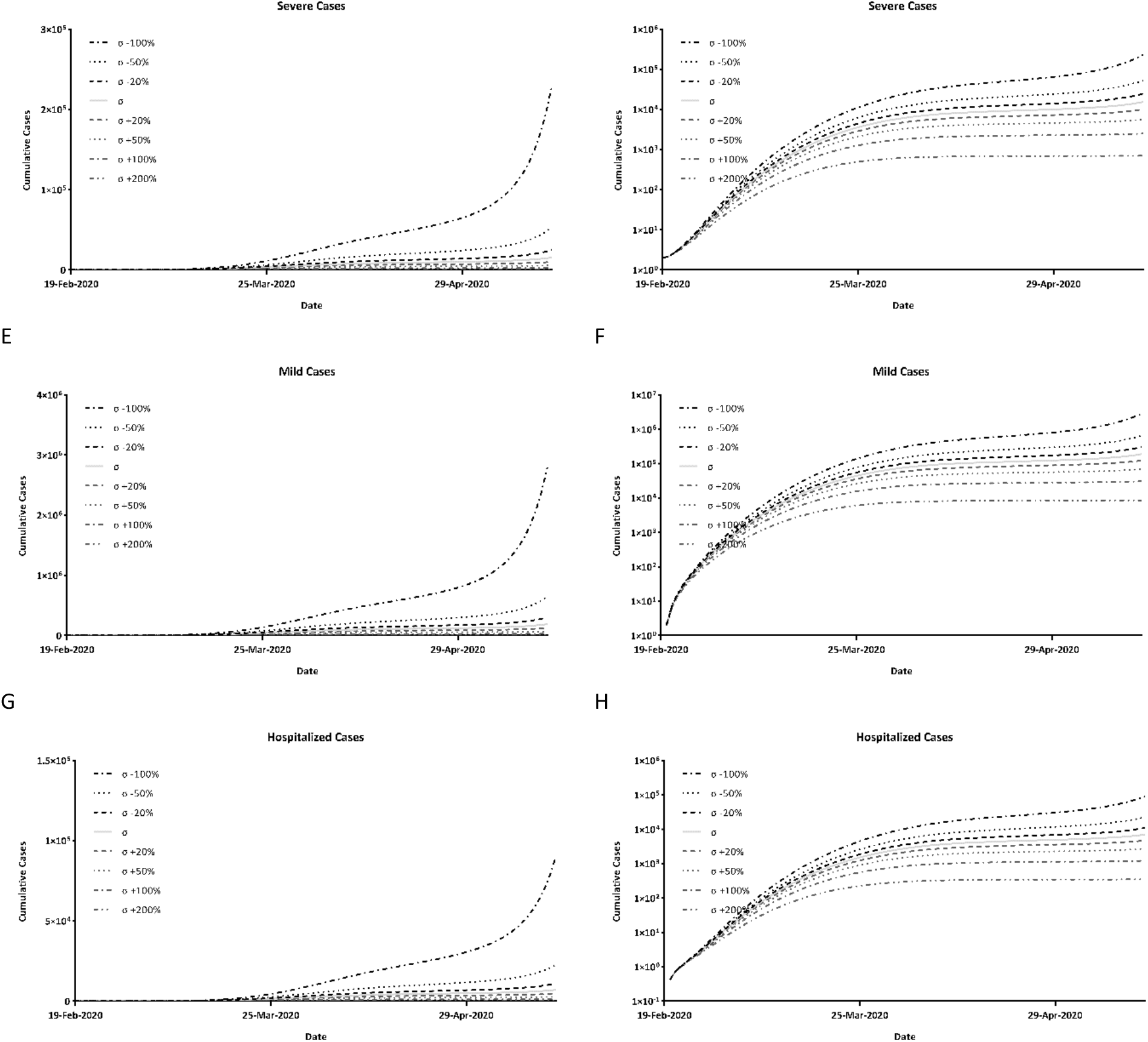
Simulation-based estimation of the cumulative number of a,b) total infected cases, c, d) severe cases, e, f) mild cases, and g, h) hospitalized cases for various scenarios. The graphs on the left are on a linear scale, and the right-side graphs are on the semi-log scale.

### 3.3 Disease-induced Fatality

Figure 7 illustrates the fatality rate induced by COVID-19 across provinces of Iran. The detailed death rates in terms of sex, as well as urban or rural population, are presented in the supplementary material (Figure S.1). By applying these rates to the total number of infected cases estimated by the SEIR model, the death tolls caused by COVID-19 were calculated. As evidenced in Figure 8, the estimated fatalities almost perfectly fitted to the reported deaths, implying the goodness of the fitted model.

**Figure 7.**
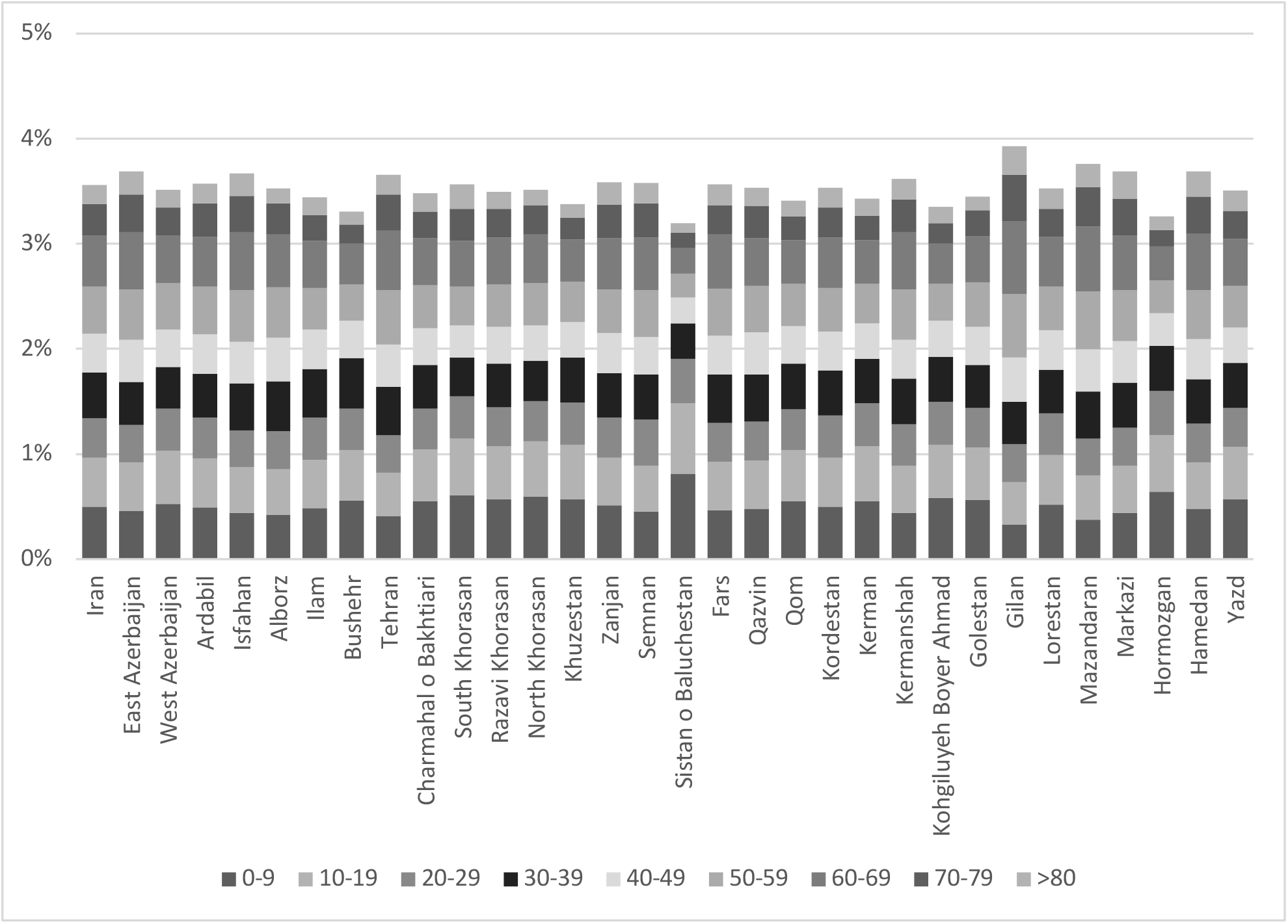
Age-adjusted disease-induced fatality rate across different provinces of Iran

**Figure 8.**
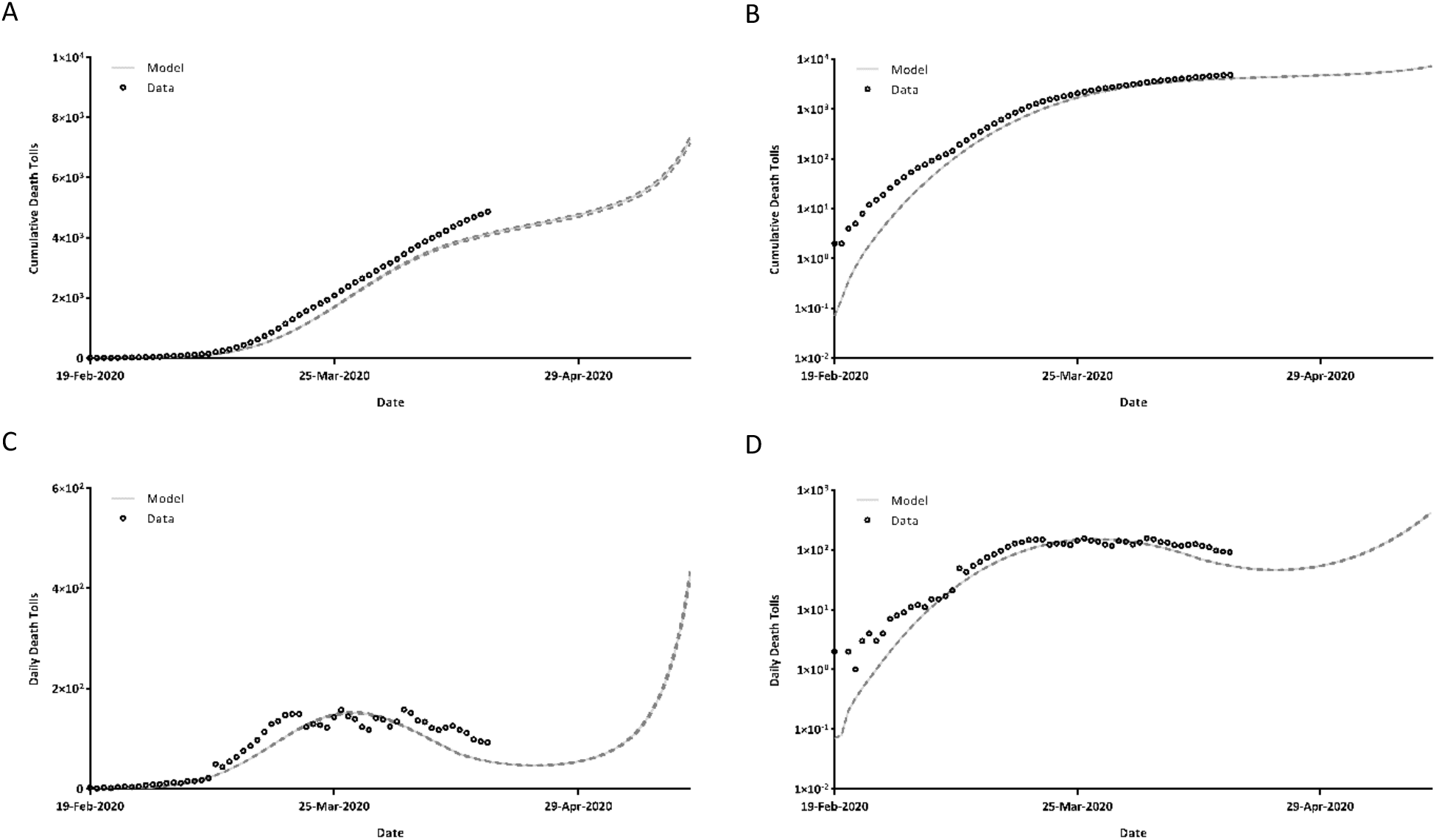
Simulation-based estimation of a, b) cumulative death tolls, and c, d) daily death tolls, based on the ground SEIR model. The graphs on the left are on a linear scale, and the right-side graphs are on the semi-log scale.

Interesting to note, the fatality rate was substantially higher for women than men (4.21% vs. 2.90%), the overall fatality rate was higher among the rural population compared to the urban population (3.62% vs 3.54%).

## Discussion

In this study, the more commonly known phenomenological (data-fitting) models and the prominent mechanistic model (SEIR) were implemented to describe the current and forthcoming trends of the COVID-19 outbreak in Iran. The data were assimilated with the official reports of the National Ministry of Health and Medical Educations to generate reliable forecasting.

Data-fitting is a beneficial tool for describing the regulation of early epidemics and making short-term forecasts; however, it does not take into account the behavioral aspects of an epidemic, such as the decisions to implement or lift the control measures. Moreover, phenomenological models are incapable of nowcasting an epidemic, e.g., giving an estimate of the real size of the epidemic at the current situation. Given in Figure 3, the GRM, RM, as well as GLGM, perfectly fitted to the data; nonetheless, there exist divergencies across their estimations of the epidemic final size, from close to 8,000 to 12,000. Hence, motivated by those mentioned above, we tended to implement a modified SEIR model to fill the gaps.

Through extending the classical SEIR model by splitting the infected class up into four subcategories, the transmission dynamics of COVID-19 in Iran was simulated. This enabled us to give an estimation of the outbreak current situation and to provide a forecast of the epidemic trend in the next month.

In Table 2, the estimated basic reproductive number at the beginning of the epidemic is compared to those specified by some investigators for Iran. The estimated *R*_*0*_ (3.659; 3.657, 3.660, 95% CI) is involved in the range of reported values (range; 2.72-6.6), suggesting the reliability of the estimations.

**Table 2.** Comparison of estimated basic reproductive number with other studies.

| Reference | Published Date | $R_0$ |
| --- | --- | --- |
| This Study | April 16, 2020 | 3.66 |
| Muniz-Rodriguez et al. (27) | April 14, 2020 | 4.4, 3.50 |
| Ahmadi et al. (5) | April 07, 2020 | 4.7 |
| Sahafizadeh et al. (28) | March 29, 2020 | 4.86 |
| Einian et al. (29) | March 28, 2020 | 6.6 |
| Ghaffarzadegan et al. (15) | March 20, 2020 | 2.72 |
| Khosravi et al. (30) | March 04, 2020 | 2.74 |

In view of the findings, around half of the infected cases were gone undetected. This, coupled with the fact that these undetected infected individuals without generating any symptoms are infectious, provides reliable evidence for quarantining the whole population.

At the early epidemic, discrepancies in some extent were observed between reported data and the SEIR model. This may be explained by the fact that the recovery rate would often increase as the time progresses after the initiation of the epidemic, whereas, in this study, a constant recovery rate was assumed throughout the time.

The model formulated in this study can be used as-is or as modified to forecast the succeeding months of the COVID-19 outbreak in Iran, and possibly by other countries, by updating the dataset used. The results in this paper can be a guide for anticipatory planning to control the COVID-19 outbreak in Iran. To have a more granular formulation of policies for decision-making, the model can be used to nowcast and forecast the epidemic values in provinces and cities.

## 4 Conclusion

Six phenomenological models were used to nowcast the epidemics in Iran by assimilating the data from the National Ministry of Health and Medical Educations. The generalized-Richards model (GRM), Richards model, generalized-logistic growth model, logistic growth model, generalized-growth model, and exponential growth model were fitted to the cumulative number of cases from 19 February 2020 to 13 April 2020. GRM, Richards model, generalized-logistic growth model, and logistic growth model projected the final epidemic size to be around 79,681, 104,482, 118,923, and 68,486 infected cases, respectively.

A modified mechanistic Susceptible-Exposed-Infectious-Recovered model was also formulated to project various scenarios, in which the parameters were calibrated based on real cumulative incidence data. It was estimated that the epidemic started with a basic reproductive number *R*_*0*_ of 3.66, then, the effective reproductive number was reduced below unity after March 27. However, it started increasing from April 11, as the quarantine measures were lifted, and it passed above unity again after April 23. The findings suggest that a second wave of outbreak may start again around April 26.

The model predicts that the total number of infected cases with COVID-19 is higher by around 50% compared to the reported numbers, implying a surge in undetected cases. Under the scenario where the epidemic will not be effectively controlled, it is estimated that around 220,000 cases would be infected by mid-May, in which 7.5% of these cases will be severely infectious, and 7,500 cases need hospitalization. Moreover, it was observed that the fatality rate was substantially higher for women (4.21%) than men (2.90%), and the overall fatality rate was higher among the rural communities (3.62%) compared to the urban (3.54%).

## Data Availability

All models are in the supplementary material. Codes can be requested from the authors.

## Acknowledgment

JFR is supported by the Abdus Salam International Centre for Theoretical Physics (ICTP) Associateship Scheme, Trieste Italy.

The authors declare no competing interests.

## 1 Phenomenological models

### 1.1 Exponential growth model

The exponential growth function is of great renown. In the dynamics of an epidemic, with the absence of any restrictions, the cumulative number of infected individuals (*C(t)*) will vary against time (*t*) in line with the following differential equation:

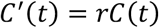

where *r* is the growth rate per time.

### 1.2 Generalized-growth model

In contrast to the assumption of the exponential growth model, after the establishment of an epidemic into a community, contacts would be highly restricted through applying control measures by the authorities. Hence, the growth of the epidemic diverges from the exponential function. Suggested by Viboud et al. (1), a decelerating factor, *p*, embedded in the exponential growth model can be used to depress the growth speed. The model is given by the following differential equation:

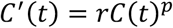

### 1.3 Logistic growth model

The logistic growth model is the known S-shaped introduced by Verhulst (2) in 1838 to model the biological population growth. The model is defined by the following equation:

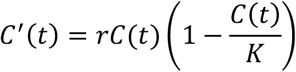

where *K* is the maximum size of the population.

### 1.4 Generalized-logistic growth model

The generalized-logistic growth model takes advantage of the decelerating factor, *p* (3), and is defined by

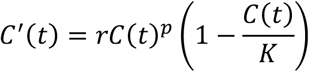

### 1.5 Richards model

The Richards model, as its name implies, formulated by Richards (4) in 1959, extends the logistic growth model by implementing a factor, *a*, to allow the model to tilt from the symmetric S-shaped. The model is defined by the following differential equation:

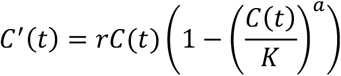

### 1.6 Generalized-Richards model

The most recently established generalized-Richards model extends the simple Richards model by introducing *p*, a parameter that decelerates the growth speed (5). The model is defined by the following differential equation:

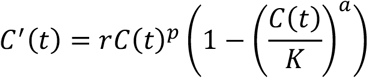

## 2 Calculation of transmission rate

The disease dynamics across the compartments were fashioned by the following system of non-linear differential equations:

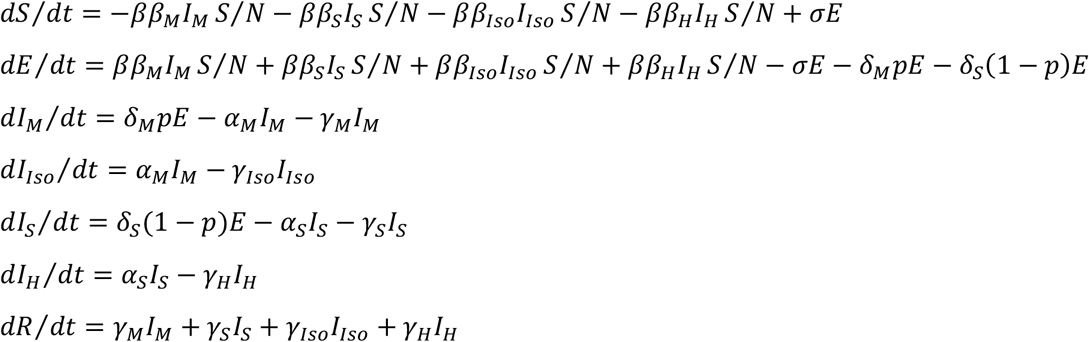

Following the proposed approach by Diekmann and coworkers (6), the next-generation matrix was formed through

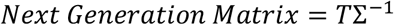

where C and Σ are respectively the transmission and transition matrix,

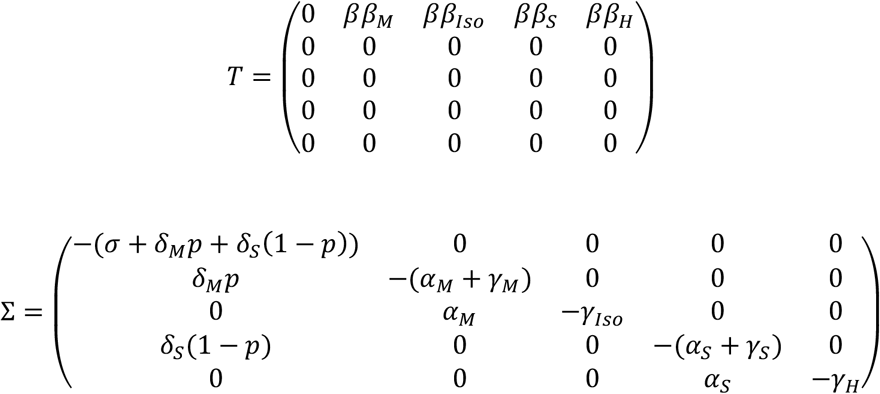

By calculating the spectral radius of the next-generation matrix, the reproduction number was derived as a function of parameters, which then, was solved for *β*.

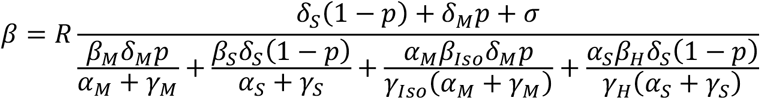

## 3 Curve Fitting

**Table S.1.** Parameter estimations for the Phenomonogical models; data were reported in median (first quartile, third quartile) form.

| Model | r | p | a | K |
| --- | --- | --- | --- | --- |
| Exponential Growth Model | 0.193 (0.193 , 0.193) | - | - | - |
| Generalized-Growth Model | 7.128 (7.029 , 7.225) | 0.538 (0.537 , 0.540) | - | - |
| Logistic Growth Model | 0.281 (0.280 , 0.281) | - | - | 68486 (68339 , 68634) |
| Generalized-Logistic Growth Model | 2.143 (2.100 , 2.190) | 0.699 (0.696 , 0.702) | - | 118923 (116933 , 121028) |
| Richards Model | 2.789 (2.277 , 3.607) | - | 0.023 (0.018 , 0.029) | 104482 (103228 , 105754) |
| Generalized-Richards Model | 3.690 (3.612 , 3.760) | 0.619 (0.616 , 0.621) | 5.284 (4.829 , 5.793) | 79681 (78733 , 80677) |

**Table S.2.** Measures of fitting accuracy for the six models.

| Model | RMSE* | MAE<sup>§</sup> | MAPE<sup>€</sup> |
| --- | --- | --- | --- |
| Exponential Growth Model | 23209 | 17537 | 81% |
| Generalized-Growth Model | 2402 | 1857 | 102% |
| Logistic Growth Model | 6336 | 4943 | 50% |
| Generalized-Logistic Growth Model | 1485 | 1172 | 22% |
| Richards Model | 2117 | 1573 | 25% |
| Generalized-Richards Model | 1010 | 810 | 44% |

\*RMSE: Root Mean Squared Error; <sup>§</sup>MAE: Mean Absolute Error; <sup>€</sup>MAPE: Mean Absolute Percentage Error

## 4 Fitted SEIR model

**Table S.3.** Estimated parameters for the SEIR model; data were reported in median (first quartile, third quartile) form.

| Parameter | Definition | Value |
| --- | --- | --- |
| $R_0$ | Basic reproductive number | 3.659 (3.657, 3.660) |
| $\beta$ | Transmission rate | - |
| $\beta_S$ | Considered as unity to scale the level of infectiveness of other infectious subgroups | 1 |
| $\beta_M$ | Relative infectiousness of mildly infected to severely infected compartment | 0.182 (0.175, 0.188) |
| $\beta_{Iso}$ | Relative infectiousness of isolated mildly infected to severely infected compartment | 0.010 (0.010, 0.010) |
| $\beta_H$ | Relative infectiousness of hospitalized to severely infected compartment | 0.150 (0.149, 0.150) |
| $\delta_M$ | Transition rate from exposed class to mildly infected subgroup (1/day) | 0.218 (0.218, 0.218) |
| $\delta_S$ | Transition rate from exposed class to severely infected subgroup (1/day) | 0.167 (0.166, 0.167) |
| $p$ | Proportion of mildly infected population to the total infected cases | 0.904 (0.904, 0.904) |
| $\alpha_M$ | Isolation rate of mildly infected cases (1/day) | 0.578 (0.577, 0.580) |
| $\alpha_S$ | Hospitalization rate of severely infected cases (1/day) | 0.262 (0.260, 0.264) |
| $\gamma_M$ | Recovery rate of mildly infected cases (1/day) | 0.310 (0.296, 0.316) |
| $\gamma_S$ | Recovery rate of severely infected cases (1/day) | 0.262 (0.261, 0.263) |
| $\gamma_{Iso}$ | Recovery rate of isolated mildly infected cases (1/day) | 0.305 (0.305, 0.306) |
| $\gamma_H$ | Recovery rate of hospitalized (1/day) | 0.256 (0.255, 0.257) |
| $T_0$ | Threshold time at which control measures were applied (day) | 7.500 (7.491, 7.507) |
| $\mu$ | Decay constant (1/day) | 0.041 (0.041, 0.041) |
| $\sigma$ | Protection rate of exposed people from being infectious (1/day) | 0.064 (0.063, 0.064) |

**Table S.4.** Measures of fitting accuracy for the SEIR model.

| RMSE* | MAE<sup>§</sup> | MAPE<sup>€</sup> |
| --- | --- | --- |
| 2772 | 1897 | 32% |

\*RMSE: Root Mean Squared Error; <sup>§</sup>MAE: Mean Absolute Error; <sup>€</sup>MAPE: Mean Absolute Percentage Error

## 5 Fatality Rates

**Figure S.1.**
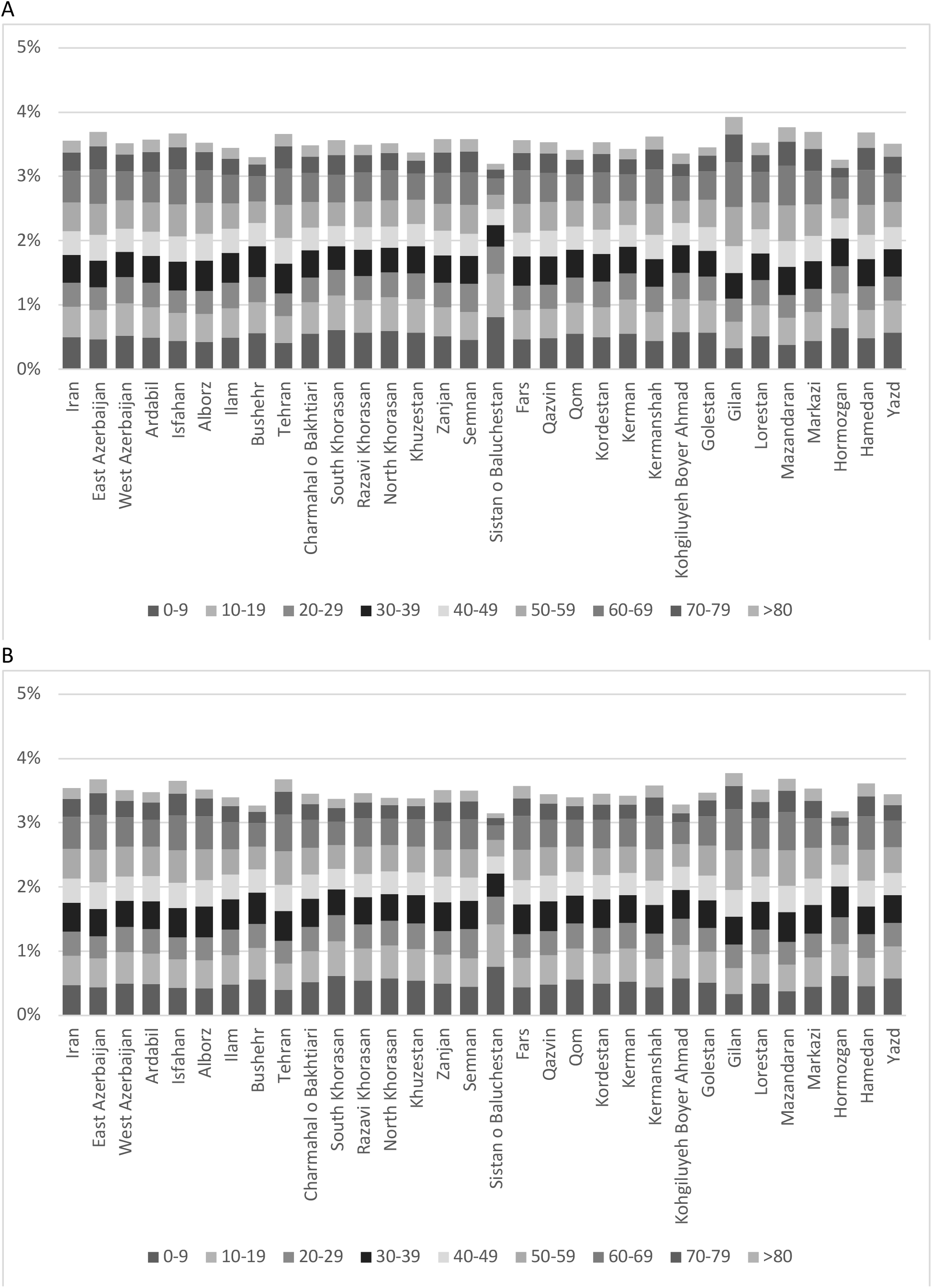
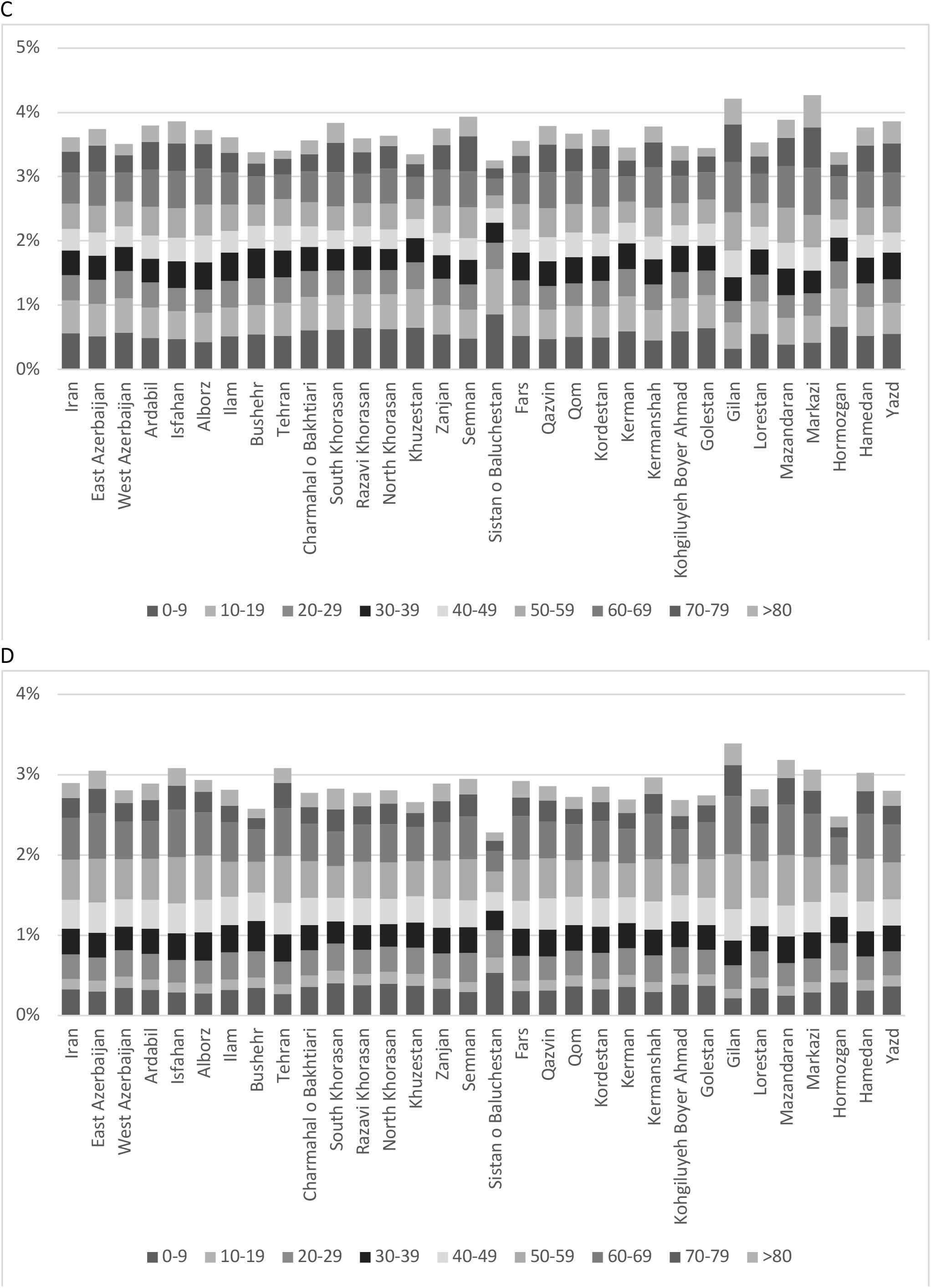
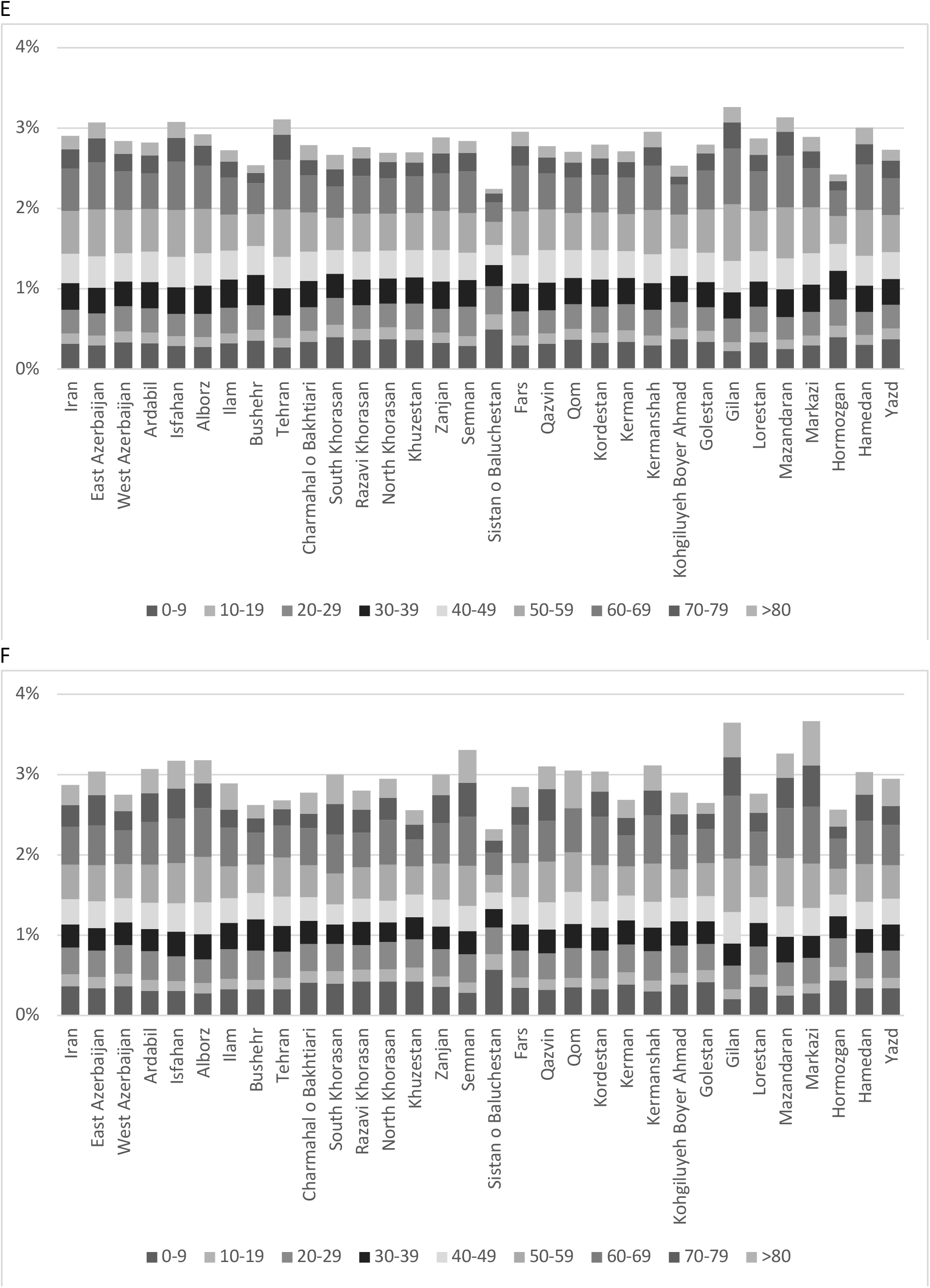
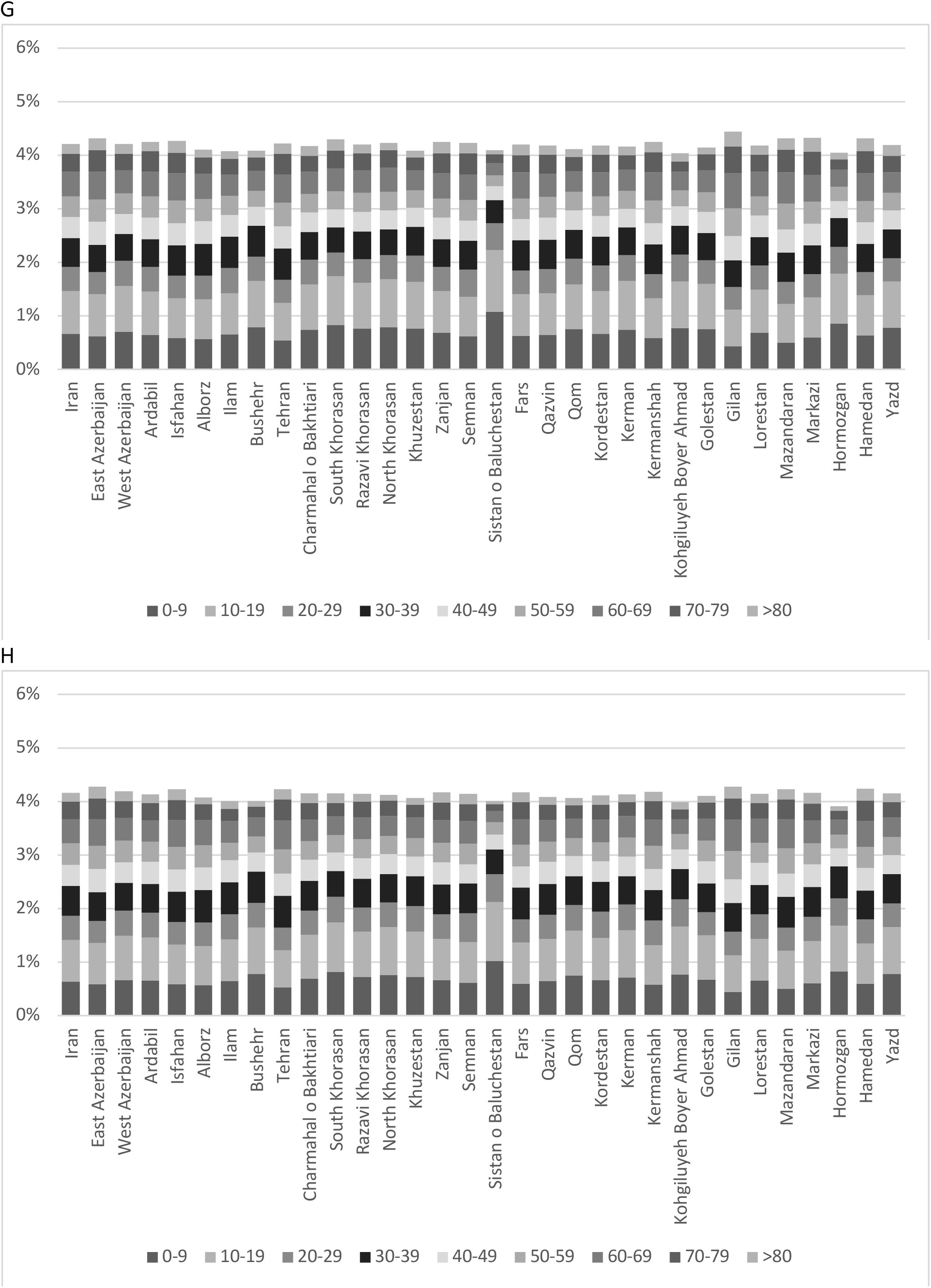
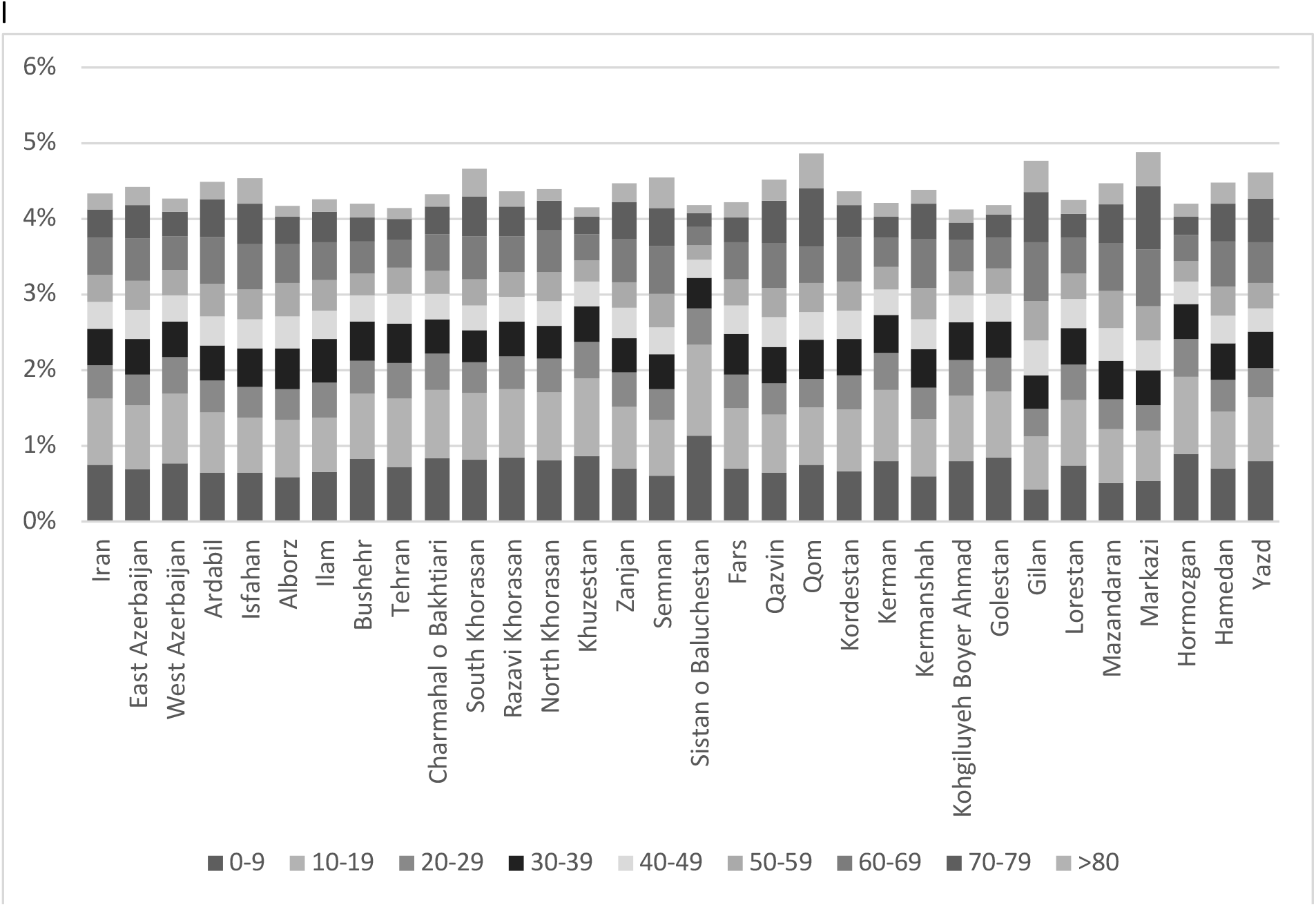
Age-adjusted fatality rate of COVID-19 across different provinces for a) overall population, b) urban population, c) rural population, d) male population, e) urban male population, f) rural male population, g) female population, h) urban female population, and i) rural female population

